# Towards uniform recognition of child abuse in the Netherlands: implementing the Screening instrument for Child Abuse and Neglect (SCAN)

**DOI:** 10.1101/2025.01.24.25321079

**Authors:** E.A.L. van den Heuvel, D.S. de Vries, B.M. de Jong- van Kempen, R. Bakx, R.F. Bos, P. van Empelen, F. Hoedeman, T.H. Kappen, I.M.B. Russel-Kampschoer, P. Puiman, M.C.M. Schouten, A.H. Teeuw, C.J. Zwaans, S.L. Nijhof, E.M. van de Putte

## Abstract

**STUDY PURPOSE:** This study investigates compliance with the Screening Instrument for Child Abuse and Neglect (SCAN screening) and the usability of SCAN screening and its follow-up tool (SCAN-FU) for SCAN-positive outcomes, in 9 Emergency Departments (EDs). It aims to address the decline in child maltreatment recognition observed over the past decade in Dutch EDs.

**METHODS:** SCAN screening and SCAN-FU were integrated into the ED-workflow and electronic health records (EHRs), supported by an implementation strategy, e-learning, policy manuals, and an awareness campaign. Compliance was evaluated by comparing pre- and post-implementation screening rates, with clinically relevant improvement defined as a ≥10% increase. Usability was assessed using the Measurement Instrument for Determinants of Innovations and semi-structured interviews. Subgroup analyses were conducted by hospital type, EHR, profession type, and years of working experience.

**RESULTS:** After implementation the average compliance rate increased from 57.5% to 70.5%, with 3 of 8 sites achieving a ≥10% improvement. Compliance rates varied by site, with academic EDs benefiting most. Compliance was influenced by EHR configurations. Usability analysis identified five facilitators (perception of responsibility, social support, self-efficacy, knowledge, and formal ratification by management) and one barrier (unsettled organisation). Users considered SCAN screening and SCAN-FU user-friendly, though perceived support differed between nurses and physicians due to role-specific factors.

**RELEVANCE:** SCAN screening can improve compliance in recognising child maltreatment, but tailored strategies are needed for further implementation. SCANs standardised approach enhances uniform data collection, enabling comparative analysis and interdisciplinary collaboration, advancing early detection of child maltreatment.

## 1. INTRODUCTION

Child maltreatment is a pervasive global issue with far-reaching consequences, affecting millions of children each year [1]. The profound impact is evident in the heightened morbidity and mortality rates among victimised children [2]. In addition to the personal harm, societal costs are substantial, with annual economic losses for adverse childhood events (including maltreatment) in the Netherlands estimated at 28.1 billion US dollars, representing 3.1% of the country’s gross domestic product [3].

Early recognition of child maltreatment is crucial to prevent further harm and reduce long-term consequences [4–6]. Yet only 29% of the European emergency departments use a screening instrument such as ESCAPE and SPUTOVAMO, which have been shown to improve detection rates [7–13]. Barriers to recognising maltreatment in emergency department include the absence of validated screening instruments, comprehensive hospital policies, and the lack of staff training [9,14,15].

Since 2013, Dutch law requires all professionals working with children to assess potential signs of maltreatment using a mandatory reporting code. Sector-specific adaptations have been developed for various contexts, including healthcare, education, and childcare. To ensure compliance, the Health Inspectorate mandates the use of a screening instrument for all children accessing hospital services [16]. When maltreatment is suspected, physicians follow the medical reporting code issued by the Royal Dutch Medical Council (KNMG), which provides a five-step framework to determine whether safety measures can be arranged, or if reporting to ‘Safe at Home’ (‘Veilig Thuis’, the Dutch centre for advice and reporting on interpersonal violence and child abuse) is necessary (figure 1, panel C)[17].

**Figure 1:**
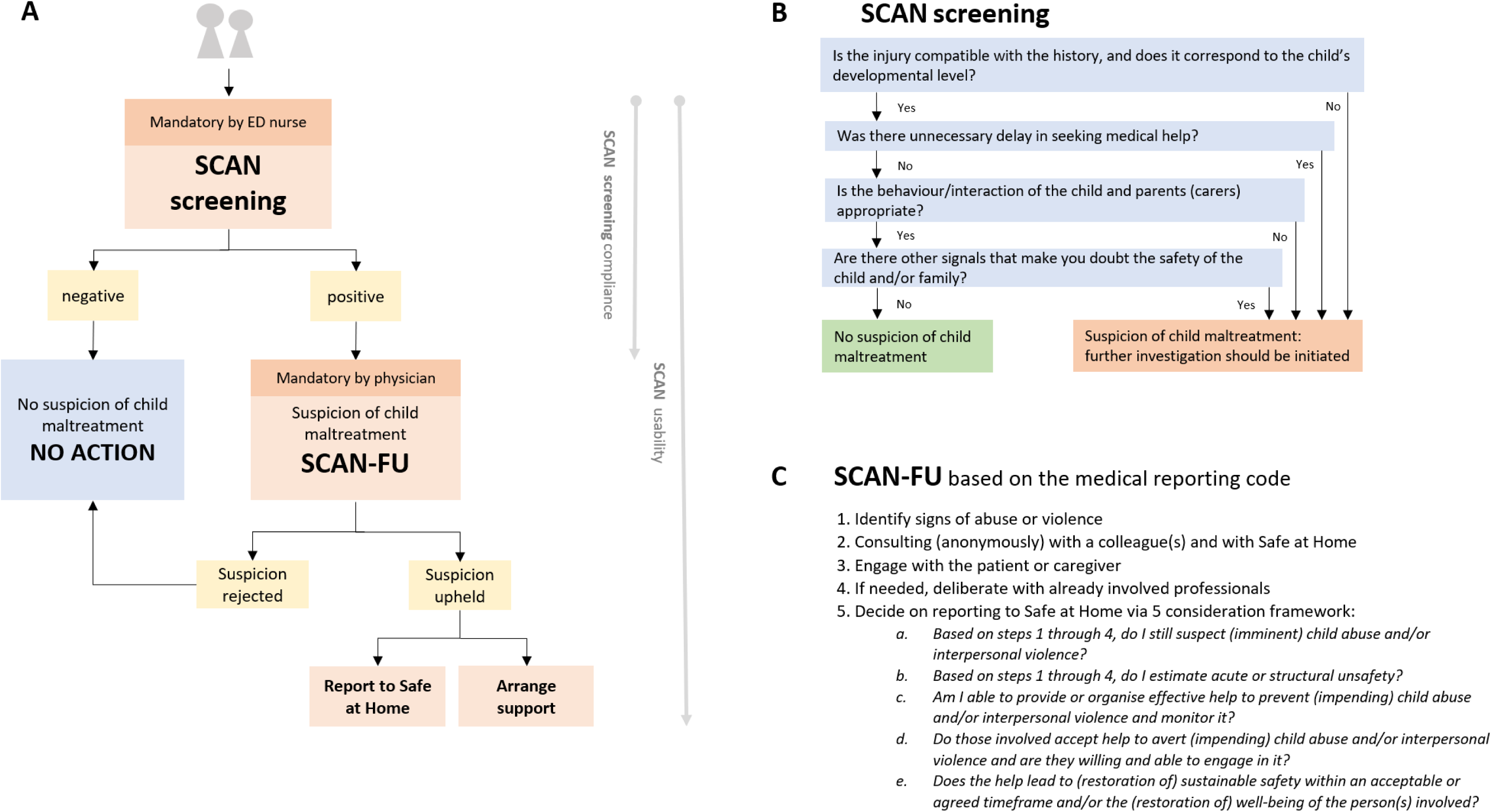
Panel A: SCAN work process within the EHR (panel A), mandatory actions in bold; work process (arrows) with spteucdifyiccsomponents. Panel B: SCAN screening [22], Panel C: Medical reporting code, 184 foundation for SCAN-FU [17].

Over the past decade, the estimated prevalence of child maltreatment in the Netherlands remained unchanged [18,19]. During this period reporting to ‘Safe at Home’ by hospital-based healthcare professionals has declined [20,21]. This suggests reduced recognition of possible maltreatment, despite mandatory staff training on child abuse recognition mandated by the Inspectorate of Health [22]. A possible contributing factor may be the lack of a validated screening instrument, resulting in variable maltreatment recognition protocols and inconsistent hospital policies. These inconsistencies hinder inter-hospital comparability, limiting opportunities for shared learning and system-wide healthcare improvement. To address this hiatus, the Screening instrument for Child Abuse and Neglect (figure 1, panel B) was developed as a uniform and validated instrument for early recognition of child maltreatment in the emergency department [23].

This study aims to evaluate the compliance with use of the SCAN screening on the recognition of possible child maltreatment in the emergency department of 9 Dutch hospitals. To optimise implementation and extend support beyond screening, additional standardised guidance has been developed to assist physicians in navigating the medical reporting code: SCAN follow-up (SCAN-FU). For the purpose of this document, the SCAN screening instrument will be referred to as the SCAN screening, and the combination of the SCAN screening and the SCAN-FU will be referred to as the SCAN. The second aim for this study is to evaluate the usability of the SCAN. The results are intended to support the nationwide implementation of the SCAN, with the overall goal of improving the early identification of child maltreatment.

## 2. METHODS

### Study design

An observational mixed-methods study was conducted. Compliance to the SCAN screening was assessed quantitatively by comparing the compliance rate with the use of a screening instrument pre- and post-implementation. A clinically relevant effect was defined as an increase of ≥ 10% of the compliance rate.

The usability of the SCAN was evaluated through a combination of data sources. This included quantitative data from the Measurement Instrument for Determinants of Innovations (MIDI) and insights gathered from in-depth semi-structured interviews. Descriptive characteristics of MIDI respondents were obtained.

To ensure broad applicability of the SCAN, a purposeful sample of hospitals was selected, including 3 academic hospitals, 4 teaching hospitals, and 2 general hospitals. These 9 hospitals, collectively operating 12 ED locations are all hospital-based and provided care for patients under 18 years (among other age groups).

The Medical Research Ethics Committee (MREC) of the University Medical Centre Utrecht classified this study as exempt according to the Medical Research Involving Human Subjects Act (MREC protocol number 20-828/C).

Patients or the public were not involved in the design, conduct, reporting, or dissemination plans of this research.

### Preparatory to SCAN implementation

Three preparatory steps to effectively implement the SCAN were designed to address known barriers and facilitators [14,15]. Firstly, to assist physicians with guidance through the medical reporting code, a standardised SCAN-follow up (SCAN-FU) tool was developed, comprising of clarifying questions per step of the reporting code (available on request).

Secondly, the SCAN was integrated into the emergency department work process and incorporated into two widely used electronic health record (EHR) systems. Within this work process (figure 1, panel A), agreements were established, assigning nurses the responsibility of the SCAN screening completion. When the SCAN screening results indicate potential child maltreatment, by one or more deviant answers (SCAN-positive), physicians are supported through the medical reporting code by SCAN-FU for further evaluation. To ensure seamless follow-up a SCAN screen-positive outcome is directly linked to the SCAN-FU within the EHR. All children (< 18 year) presented at the ED were eligible for the use of SCAN, there were no exclusion criteria.

Thirdly, to support the implementation, several resources were developed: 1) a concise e-learning to educate professionals on the SCAN work process; 2) a policy manual outlining the process including the SCAN screening and the SCAN-FU, communication strategies and legal guidelines (available on request); and 3) awareness campaign materials to prepare the staff for the SCAN implementation. All resources were provided to participating hospitals at no cost.

### Implementation strategy

Before implementation hospitals were required to fulfil specific inclusion criteria: (1) appoint a clinical ambassador, (2) integrate the SCAN into the EHR system, and (3) provide consent for data collection.

The clinical ambassador determined the start date for implementation. In collaboration with the research team, the SCAN implementation strategy was customised. The clinical ambassador received tailored guidance on critical decisions, including configuring the EHR system (workflow interruptions and system notifications), selecting colleagues for mandatory training delivered through the e-learning (at minimum: emergency department staff and paediatricians), and allocating resources for awareness campaigns. The research team provided continuous support throughout the implementation process to ensure uniform and effective execution. A more detailed description of the implementation strategy can be found in Supplement A.

### Data collection

To assess the effect of the SCAN screening in terms of the compliance with the use of a screening instrument, data collection began three months before implementation and continued for at least one year. Aggregated data were collected from each participating hospital, including the total number of emergency department visits by children under 18 years, the number of children screened (using any instrument), and the number of screening-positives.

To evaluate the usability of the SCAN, a multi-phase approach was used. First, SCAN users identified factors influencing implementation in their hospital. The clinical ambassador at each site selected a broad group of SCAN users (a minimum of 30-50, depending on the hospital size), who received the MIDI-questionnaire via email at least three months post-implementation, and up to two reminders were sent. Respondents answered anonymously and were asked to provide background information, including current profession, number of years of work experience, and experience with child abuse screening instruments and reporting to ‘Safe at Home’.

Subsequently, semi-structured in-depth interviews were conducted with a smaller representative sample of SCAN users per hospital, including a minimum of a paediatrician, an emergency department physician and an emergency department nurse. These interviews, conducted at least two months after the MIDI-questionnaire, explored the facilitators and barriers in greater detail. Using a hybrid format via Microsoft Teams, the interviews followed guided questions informed by the hospital-specific MIDI results.

SCAN implementation took place sequentially between May 2021 and March 2022.

### MIDI-questionnaire

The questionnaire used was adapted from the Measurement Instrument for Determinants of Innovation (MIDI) [24], an evidence-based framework that outlines 29 determinants influencing healthcare innovation implementation. These determinants were organised into four domains: (1) the innovation, (2) the user, (3) the organisation, and (4) the socio-political context.

From the original 29 MIDI determinants, the research team identified 17 as potentially relevant, (of which 3 included a sub question). These determinants were adapted to reflect the context of the SCAN implementation, in accordance with instructions of the MIDI questionnaire use. The questionnaire consisted of items rated either on a 5-point Likert scale (n=18), ranging from “*totally disagree*” (1) to “*totally agree*” (5), or binary response options (n=2) (table 1).

**Table 1:**
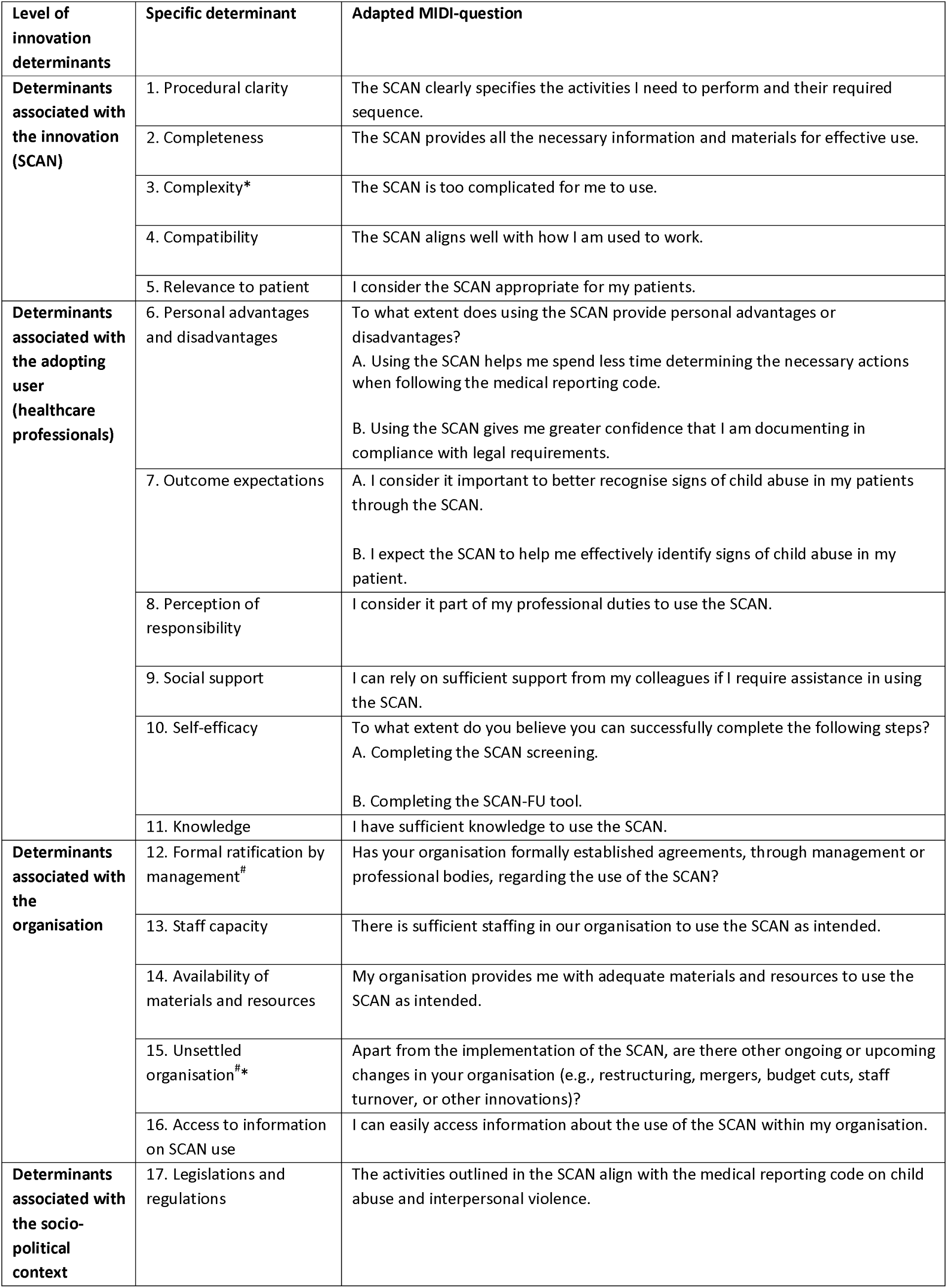
MIDI determinants adjusted for SCAN implementation. *Reverse scale, ^#^binary question.

| Level of innovation determinants | Specific determinant | Adapted MIDI-question |
| --- | --- | --- |
| **Determinants associated with the innovation (SCAN)** | 1. Procedural clarity | The SCAN clearly specifies the activities I need to perform and their required sequence. |
|  | 2. Completeness | The SCAN provides all the necessary information and materials for effective use. |
|  | 3. Complexity* | The SCAN is too complicated for me to use. |
|  | 4. Compatibility | The SCAN aligns well with how I am used to work. |
|  | 5. Relevance to patient | I consider the SCAN appropriate for my patients. |
| **Determinants associated with the adopting user (healthcare professionals)** | 6. Personal advantages and disadvantages | To what extent does using the SCAN provide personal advantages or disadvantages?<br>A. Using the SCAN helps me spend less time determining the necessary actions when following the medical reporting code.<br>B. Using the SCAN gives me greater confidence that I am documenting in compliance with legal requirements. |
|  | 7. Outcome expectations | A. I consider it important to better recognise signs of child abuse in my patients through the SCAN.<br>B. I expect the SCAN to help me effectively identify signs of child abuse in my patient. |
|  | 8. Perception of responsibility | I consider it part of my professional duties to use the SCAN. |
|  | 9. Social support | I can rely on sufficient support from my colleagues if I require assistance in using the SCAN. |
|  | 10. Self-efficacy | To what extent do you believe you can successfully complete the following steps?<br>A. Completing the SCAN screening.<br>B. Completing the SCAN-FU tool. |
|  | 11. Knowledge | I have sufficient knowledge to use the SCAN. |
| **Determinants associated with the organisation** | 12. Formal ratification by management<sup>#</sup> | Has your organisation formally established agreements, through management or professional bodies, regarding the use of the SCAN? |
|  | 13. Staff capacity | There is sufficient staffing in our organisation to use the SCAN as intended. |
|  | 14. Availability of materials and resources | My organisation provides me with adequate materials and resources to use the SCAN as intended. |
|  | 15. Unsettled organisation<sup>#*</sup> | Apart from the implementation of the SCAN, are there other ongoing or upcoming changes in your organisation (e.g., restructuring, mergers, budget cuts, staff turnover, or other innovations)? |
|  | 16. Access to information on SCAN use | I can easily access information about the use of the SCAN within my organisation. |
| **Determinants associated with the socio-political context** | 17. Legislations and regulations | The activities outlined in the SCAN align with the medical reporting code on child abuse and interpersonal violence. |

To ensure the appropriateness of the selected determinants, a preliminary in-depth interview was conducted, two months after the SCAN implementation in the first hospital but before distributing the MIDI-questionnaire. This interview involved a representative sample of users and aimed to identify facilitators and barriers to SCAN implementation. The findings closely aligned with the selected MIDI determinants, affirming their relevance. Consequently, no further modifications to the MIDI determinants were made. The MIDI-questionnaire was hereafter analysed prior to conducting the semi-structured interviews.

### Data analysis

IBM SPSS Statistics for Windows (version 29.0.1.0, Armonk, NY, USA; IBM Corp.) was used for quantitative data analysis. Descriptive statistics were applied to assess the compliance to screening and usability of SCAN from the MIDI questionnaire (including mean, interquartile range, percentages, standard deviation, Chi-square tests, and ANOVA).

Consistent with prior research, MIDI determinants phrased positively were classified as facilitators if ≥80% of participants responded with “*agree*” or “*strongly agree*” [25]. Conversely, determinants were categorized as barriers if ≥20% of participants responded with “ *disagree*” or “*totally disagree*.” The reliability of the SCAN-adapted MIDI-questionnaire was evaluated using Cronbach’s alpha, which indicated good internal consistency (α = 0.85).

Prespecified subgroup analyses examined differences in compliance rate and usability by hospital and EHR type. Subgroup analysis for useability were furthermore performed by work experience (early career [<5 years], mid-career [6–15 years], experienced [>15 years]) and type of profession (nurse, physician) to account for task-specific responsibilities within the SCAN workflow. In accordance with standard practice, a p-value <0.05 was used to define statistical significance in subgroup analysis.

## RESULTS

All participating hospitals fulfilled the inclusion criteria and were able to implement the SCAN within the study period.

### Compliance with SCAN screening

Prior to the SCAN implementation all hospitals employed a child maltreatment screening instrument, with an average compliance rate of 57.5% (range: 11%–100%). Post-implementation, this increased to 70.8% (range: 24%–100%) (table 2). The proportion of children visiting the emergency department who screened positive remained consistent, at 2.5% pre-implementation (range: 0.9%–4.0%), and 2.3% post-implementation (range: 0.7%–6.5%).

**Table 2:** ED presentation of children pre- (left) and post-implementation. Hospital I was not able to retrieve the pre-implementation data because the SCAN implementation started simultaneously with a go-live of a new EHR. Hospital^1^ = academic, hospital^2^ = teaching, hospital^3^= general. EHR A-D = Chipsoft, E-I= EPIC.

|  | PRE SCAN Screening |  |  |  |  |  | SCAN screening |  |  |  |  |  |
| --- | --- | --- | --- | --- | --- | --- | --- | --- | --- | --- | --- | --- |
|  | Children at ED | Screened children | Screened children | Screen positive | Screen positive / children at ED | Period | Children at ED | Screened children | Screened children | Screen positive | Screen positive / children at ED | Period |
| Hospital | Total (n) | Total (n) | % | Total (n) | % | Months | Total (n) | Total (n) | % | Total (n) | % | Months |
| A<sup>3</sup> | 618 | 618 | 100 | 25 | 4.0 | 3 | 6564 | 5945 | 90.6 | 211 | 3.2 | 23 |
| B<sup>1</sup> | 1115 | 131 | 11.8 | 33 | 2.9 | 3 | 5953 | 3154 | 53 | 210 | 3.5 | 20 |
| C<sup>1</sup> | 811 | 799 | 98.5 | 35 | 4.3 | 3 | 3834 | 3834 | 100 | 250 | 6.5 | 16 |
| D<sup>2</sup> | 1115 | 597 | 53.5 | 17 | 1.5 | 3 | 7459 | 5059 | 67.8 | 118 | 1.5 | 15 |
| E<sup>2</sup> | 1663 | 497 | 29.9 | 15 | 0.9 | 3 | 14443 | 3559 | 24.6 | 170 | 1.2 | 13 |
| F<sup>1</sup> | 1565 | 402 | 25.7 | 26 | 1.7 | 3 | 15621 | 11008 | 70.5 | 244 | 1.5 | 21 |
| G<sup>2</sup> | 2091 | 1376 | 65.8 | 50 | 2.3 | 3 | 22199 | 14073 | 63.4 | 247 | 1.1 | 22 |
| H<sup>2</sup> | 772 | 576 | 74.6 | 21 | 2.7 | 3 | 6014 | 4258 | 70.8 | 41 | 0.7 | 14 |
| I<sup>3</sup> |  |  |  |  |  |  | 2266 | 2121 | 93.6 | 26 | 1.5 | 12 |
| **Total** | **9750** | **4996** |  | **222** |  | **24** | **84353** | **53011** |  | **1517** |  | **156** |
| **Average** |  |  | **57.5** |  | **2.5** | **3** |  |  | **70.5** |  | **2.3** | **17** |

Compliance rates increased in 4 of the 8 hospitals, with 3 achieving gains exceeding >10%. In the other hospitals, compliance rates decreased (range 2.4% to 9.4%). At hospital A, a reorganization of emergency care services during the COVID-19 pandemic led to the erroneous inclusion of children assessed outside the emergency department as emergency department cases in the post-implementation data, contributing to a 9.4% compliance rate decrease. Despite this, hospital A maintained a higher compliance rate than most emergency departments due to its strong pre-implementation performance.

Subgroup analysis of SCAN screening showed that there were significant differences in compliance rates between academic and non-academic EDs pre-implementation (45% vs 65%, p<0.001). After implementation the difference reversed (75% vs 63%, p<0.001) with academic emergency departments outperforming non-academic emergency departments.

Differences in compliance rate were also observed between different EHR systems. Three of 4 Chipsoft hospitals increased compliance rates, compared to 1 of 4 EPIC hospitals, though this was not statistically significant (p = 0.1). Hospitals A–D implemented a “stop function” requiring emergency department-nurses to complete the SCAN screening before finalizing the consultations. Variations in default settings within Chipsoft affected compliance levels, with hospital C achieving 100% compliance under optimized conditions. In contrast, hospitals E–I lacked this functionality in their EHR systems.

During the in-depth interviews, participants reported that despite the potential “stop function” hospitals hesitated to implement it due to concerns about delaying the transfer of critically ill patients from the emergency department to the Intensive Care Unit or Operating Room if the SCAN screening was incomplete. Other barriers expressed were workload pressure, leading to the SCAN screening being frequently overlooked or omitted. Hospital E attributed low compliance to high staff turnover and infrequent training on recognising abuse, limiting the integration of SCAN into routine practice for many professionals.

SCAN users acknowledged the benefits of SCAN screening, particularly due to improved workflow agreements. One nurse working in hospital G explained, “*Although the compliance rate did not increase, the workflow in the emergency department changed significantly. Prior to implementation, the child abuse screening instrument was often completed retrospectively by members of the child abuse team, mostly after the child had already left the emergency department. With the SCAN workflow agreements, this responsibility lies explicitly with the treating nurse who actually sees the patient. This is more accurate and also enabled the child abuse team to focus more on complex cases rather than on completing an instrument*.”

### Usability of SCAN

The MIDI questionnaires were administered on average five months after the implementation of the SCAN (range: 2–9 months). A response rate could not be determined due to the distribution method, as local ambassadors distributed the questionnaire link via email to (groups of) SCAN users or included it in departmental newsletters. Of the 259 healthcare professionals who participated in the questionnaire, 173 completed the questionnaire in its entirety. Characteristics of MIDI-responders are shown in table 3.

**Table 3.** Characteristics of healthcare providers participating in MIDI questionnaire in nine hospitals PM: Other healthcare professionals include physician assistants/nurse practitioners, medical secretaries and PhD-student.

|  | Physician<br>(n) | Nurse<br>(n) | Other<br>(n) | Total<br>(n, %) |
| --- | --- | --- | --- | --- |
| **Hospital type** |  |  |  |  |
| Academic (n=3) | 43 | 32 | 7 | 82 (32) |
| Teaching (n=4) | 34 | 84 | 14 | 132 (52) |
| General (n=2) | 7 | 29 | 4 | 40 (16) |
| **Total** | **84** | **145** | **25** | **254 (100)** |
| **Work experience** |  |  |  |  |
| Years in current job (mean, SD) | 10 ($\pm 7.3$) | 14 ($\pm 10.3$) | 11 ($\pm 9.3$) | |
| Experience with a child abuse screening instrument | 84 | 143 | 23 | 250 (98.4%) |
| Experience with reporting to Safe at Home | 84 | 141 | 23 | 248 (97.6%) |

In-depth interviews were conducted at on average 2.5 months after the administration of the MIDI-questionnaire (range: 1–5 months). A total of 46 professionals participated, with an average of five participants per hospital.

### Facilitators

MIDI questionnaires found five facilitators of impact when implementing the SCAN (table 4).

**Table 4:** MIDI results of implementation facilitators and barriers of the SCAN. *Reverse scale, ^#^binary question.

| MIDI item | Respondents<br>(N) | Median<br>(IQR) | Disagree /<br>totally<br>disagree<br>(%) | Neutral<br>(%) | Agree /<br>totally<br>agree<br>(%) |
| --- | --- | --- | --- | --- | --- |
| 1.Procedural clarity | 213 | 4 (4-4) | 5.6 | 16.9 | 77.5 |
| 2.Completeness | 205 | 4 (3-4) | 6.3 | 26.3 | 67.3 |
| 3.Complexity* | 204 | 4 (3-4) | 65.2 | 25.5 | 9.8 |
| 4.Compatibility | 203 | 4 (3-4) | 8.9 | 27.1 | 64 |
| 5.Relevance to patient | 200 | 4 (4-4) | 4.5 | 19 | 76.5 |
| 6a. Personal advantages/disadvantages -<br>actions | 188 | 3.5 (3-4) | 13.8 | 36.2 | 50 |
| 6b. Personal advantages/disadvantages -<br>documentation | 186 | 3 (3-4) | 9.7 | 41.9 | 48.4 |
| 7a. Outcome expectations - recognition | 184 | 4 (3.25-4) | 4.8 | 20.1 | 75 |
| 7b. Outcome expectations – expectance | 181 | 3 (3-4) | 9.4 | 55.2 | 35.4 |
| 8.Perception of responsibility | 181 | 4 (4-5) | 2.3 | 7.7 | 90.1 |
| 9.Social support | 181 | 4 (4-4) | 2.2 | 14.4 | 83.4 |
| 10a.Self-efficacy – SCAN screening | 180 | 4 (4-4) | 0.6 | 8.9 | 90.5 |
| 10b.Self-efficacy – SCAN follow-up | 180 | 4 (3-4) | 4.4 | 29.4 | 66.3 |
| 11.Knowledge | 179 | 4 (4-4) | 3.4 | 16.2 | 80.4 |
| 12.Formal ratification by management<sup>#</sup> | 177 | 2 (2-2) | 13 |  | 87 |
| 13.Staff capacity | 175 | 4 (3-4) | 6.3 | 28.6 | 65.1 |
| 14.Availability of materials and resources | 175 | 4 (3-4) | 3.5 | 25.7 | 70.9 |
| 15.Unsettled organisation<sup>**</sup> | 175 | 2 (1-2) | 44.6 |  | 55.4 |
| 16.Access to information on SCAN tools use | 175 | 4 (4-4) | 2.3 | 20.6 | 77.1 |
| 17.Legislations and regulations | 173 | 4 (4-4) | 2.3 | 19.1 | 78.6 |

**Table 5:**
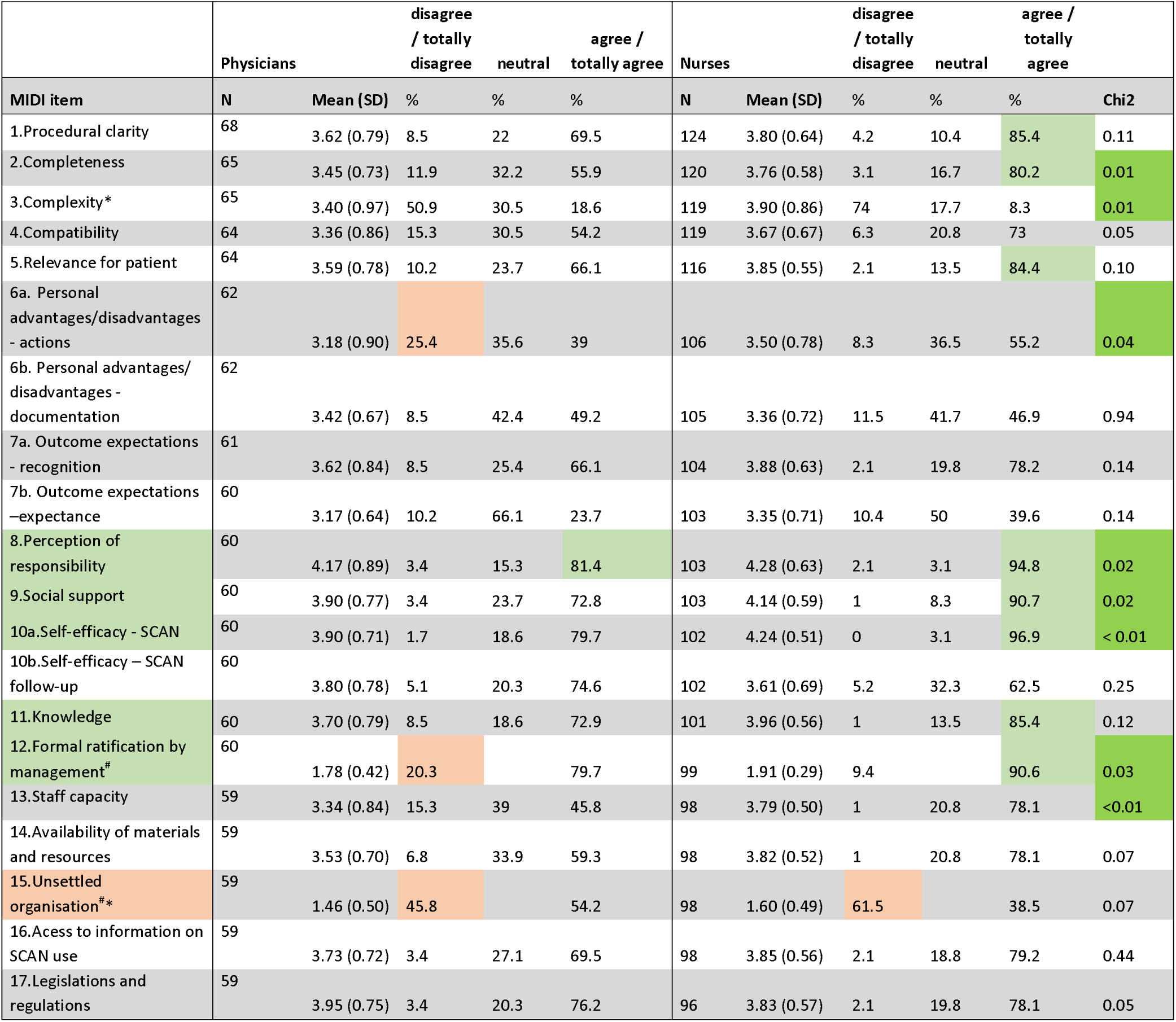
Subgroup analysis MIDI determinants – profession type (left: physicians)

Four of these were in the domain of the user and one in the domain of the organisation. Participants provided the following descriptions during the interview:

#### 1. Perception of responsibility

Ninety percent of respondents acknowledged that recognising child maltreatment is a fundamental aspect of their professional duties. They emphasised their pivotal role in identifying potential unsafe situations, noting that their focused yet brief interactions with children and parents at the emergency department provides critical opportunities for recognition. As one respondent stated, “*The SCAN screening strengthens my ability as a nurse to effectively recognise signs of child abuse. It helps me articulate concerns, enabling clearer communication with the physician”*.

#### 2. Social support

Eighty-three percent of respondents reported that they can rely on their colleagues for support when completing the SCAN. Many respondents remark; “*In the emergency department, an experienced colleague is available almost 24/7, providing easy access for consultation when I have doubts about signs of child abuse*.”

#### 3. Self-efficacy SCAN screening

Over 90% of respondents reported that they are able to successfully complete the SCAN screening, describing it as brief, clear and user-friendly. As one respondent noted: “*The SCAN screening is highly practical and represents an improvement over the SPUTVAMO* [pre-implementation used screening instrument]”.

#### 4. Knowledge

Eighty percent of respondents say they have sufficient knowledge to work with SCAN. They highlighted the availability of adequate resources, such as the manual and e-learning. However, during the interviews variability in knowledge among professionals was identified as a significant challenge, potentially resulting in interprofessional discrepancies and missed signs of child maltreatment. Continuous training was repeatedly emphasised as critical to improve detection of possible signs of maltreatment, as illustrated by one respondent: “*When you are not on duty for a while, the information tends to fade, so maintaining continuous attention to training is essential.”*

#### 5. Formal ratification by management

Eighty-seven percent of respondents reported receiving support from their management, primarily through formal working agreements regarding the use of the SCAN. One respondent remarked: “*It’s very clear that the SCAN screening is mandatory and that it must be completed by default. Agreements have been made about this, and we are reminded of it verbally*.”

Respondents from hospitals without prior formal agreements highlighted the significant advantages of implementing default SCAN screening and workflow for all children within the EHR. They reported that this approach increased compliance rates by addressing inconsistent practices: previously, the screening instrument was completed by either the physicians or the nurses, and often not at all due to assumptions that another professional would handle it.

### Barrier

One key barrier identified was organisational instability. Forty-four percent of respondents reported that various organisational factors adversely affected the implementation of the SCAN. The most frequently cited was the COVID-19 pandemic, as the SCAN was introduced during the height of the crisis. Respondents emphasized that the high workload and uncertainty associated with the pandemic reduced their readiness for change and willingness to engage in learning activities.

Other organisational factors mentioned included staff shortages, high staff turnover with insufficient time for comprehensive onboarding, hospital or site mergers and reorganizations, and changing EHR system. One respondent summarized: “*Turbulence in the emergency department occasionally complicates the completion of the SCAN screening. This turbulence is not only related to workload but also to structural changes within the ED. These factors redirected time and attention away from the SCAN*.”

### Subgroup analysis

#### Profession type

Subgroup analysis of the MIDI questionnaire by profession type identified three additional facilitators for nurses (all in the domain of the SCAN), and two additional barriers for physicians (one in the user domain and the other in de organisation domain).

Perceptions of personal advantages and disadvantages (sub question “actions”) differed between nurses and physicians: A nurse stated: “*Before the SCAN, we had no support in navigating the reporting code. I still probably won’t do it independently, but now, when I have to, I know how*”. In contrast a physician noted: “*I don’t feel more confident documenting in a legally correct manner. It remains a judgment call, as every case is different. You can never completely eliminate this uncertainty in the medical reporting code*”.

These differences are caused by distinct roles and responsibilities: nurses routinely engage with SCAN screening, while physicians interact less frequently with the SCAN-FU tool. Physicians cited the extensive medical reporting code and time-consuming nature of familiarizing themselves with the SCAN-FU tool as barriers. However, when physicians gained experience with SCAN-FU, they noted that SCAN-FU made them more intuitive and supportive compared to pre-implementation practices. For example by the standardised SCAN-FU documentation, which facilitates locating the mandatory reporting steps within the EHR, enabling appropriate actions regardless of changes in the responsible physician during the process. A barrier of the SCAN-FU tool was the increased administrative burden, challenging its integration into clinical workflows.

The determinant of completeness also highlights the differences in responsibilities within the workflow and is furthermore reflected in significant variations across self-efficacy and formal ratification by management.

#### Hospital type

Subgroup analysis across different types of hospitals identified one clinically significant difference. Professionals in teaching hospitals were significantly more likely to perceive the SCAN as compatible with the medical reporting code (detailed tables in Supplement B). However, the in-depth interviews did not provide an explanation for this observed difference.

#### Working experience

The subgroup analysis of working experience indicated no clinically significant differences between professionals with different levels of working experience (detailed in Supplement B).

## DISCUSSION

The SCAN was successfully implemented in 9 Dutch emergency departments, achieving clinically significant increased compliance rates in 3 of the 8 hospitals. SCANs usability was good, as indicated by the MIDI identifying mostly facilitators, and as supported by the findings in the interviews. SCAN implementation may improve the likelihood with which potential child maltreatment can be excluded in children presenting to the ED.

Although all hospitals aimed to use SCAN screening for all children visiting the emergency department, this goal was not achieved in all. SCAN screening was most effective in improving compliance rates in academic emergency departments, whom had a low compliance rate prior to implementation, while compliance rates in non-academic emergency departments remained largely unchanged. The MIDI questionnaire and in-depth interviews did not identify specific explanations for these differences. Variations in EHR configurations appear to influence compliance, with 100% compliance achievable under optimised conditions. Future SCAN implementation efforts should focus on optimising EHR configurations.

SCAN demonstrated good usability and adequate support, likely due to pre-implementation alignment. SCAN is user-friendly and enhances interdisciplinary collaboration. Key facilitators were identified in the user domain, crucial for success as implementation depends on behavioural changes among healthcare professionals. Organisational turbulence, inherent to the emergency department, emerged as barrier. The SCAN screening, validated for use in the emergency department, may warrant expansion to inpatient and outpatient settings, as neglect is the most prevalent form of child maltreatment, with presenting symptoms and prevalence rates likely comparable across these settings. Nurses reported greater benefits from SCAN implementation than physicians, likely reflecting differences in roles and responsibilities. The SCAN-FU tool, being more extensive than SCAN screening, is less frequently used and requires case-specific customisation, limiting its perceived utility. Further research is needed to optimise its alignment with the mandatory medical reporting code, and enhanced training for physicians may improve its implementation.

This study’s strengths include a representative sample of Dutch emergency departments, a post-implementation follow-up of at least one year, and a significant number of MIDI respondents, enhancing the generalisability of the findings. The average compliance rate of 70.8% of children presenting at emergency departments aligns with prior studies [26–28], as does the percentage of screen-positive cases (1.8–2.5%) [26–29]. Considering the low baseline compliance rates for recognising potential child maltreatment prior to implementation, further efforts to expand the national adoption of SCAN are warranted. Its potential to enhance compliance rates, together with the standardised workflow it establishes, offers significant opportunities for data exchange and collaborative learning. However, significant variability in compliance rates across participating hospitals highlights the need for tailored implementation strategies. Increased compliance is only meaningful if SCAN screening and SCAN-FU are completed with careful knowledge and consideration, emphasising the need for continuous training on the topic. A limitation of this study was the short pre-implementation period and the use of aggregated pre- and post-implementation data, which prevented the analysis of trends in compliance rates per hospital and percentage screen-positive by interrupted time series analysis.

Unified data collection through SCAN simplifies hospitals’ ability to evaluate their performance in recognising child maltreatment by standardising documentation within the EHR. On a national level, such data allow for comprehensive trend analyses, offering valuable insights to optimise hospital policies. Recent research underscores the importance of data integration in improving these strategies [29–31]. In the near future SCAN should also facilitate seamless data linkage from hospitals with ‘Safe at Home’, enabling comprehensive evaluation of the entire care chain. This marks a transformative step in reducing the harmful impact of child maltreatment and enhancing the effectiveness of multidisciplinary care strategies.

## Data Availability

All data produced in the present work are contained in the manuscript

## Footnotes

### AUTHORS CONTRIBUTION

EP designed the study, together with DS, BJK, RBa, RBo, RF, IRK, PP, MCS, AT this was conceptualised, and the SCAN-FU was developed. EH and EP developed assisting implementation resources. EH, DS, BJK, RBo, FH, IRK, PP, AT, IW and CZ were responsible for local SCAN implementation and data collection (clinical ambassadors). EH carried out data analysis, TK, SL and EP supervised the analysis interpreted the data and reviewed the manuscript. All authors commented on the manuscript and agreed on the final version.

## ACKNOWLEDGEMENT

We thank M. Scheepers and K. Sijstermans (OLVG Hospital, Amsterdam), and I. Westerbeek (Amstelland Hospital, Amstelveen) for their contributions as clinical ambassadors, and Nina Onkenhout for support with the analyses

## PATIENT CONSENT FORM

Not applicable

## DATA AVAILIBILITY STATEMENT

All data relevant to the study are included in the article or uploaded as supplementary information.

## Abbreviations

ED: Emergency Department
EHR: Electronic Health Record
IQR: Interquartile range
MIDI: Measurement Instrument for Determinants of Innovation
SCAN: Screening instrument for Child Abuse and Neglect
SCAN-FU tool: Screening instrument for Child Abuse and Neglect follow-up tool

