## Supplement A for "Towards uniform recognition of child abuse in the Netherlands: implementing the Screening instrument for Child Abuse and Neglect (SCAN)"

**Implementation Strategy**

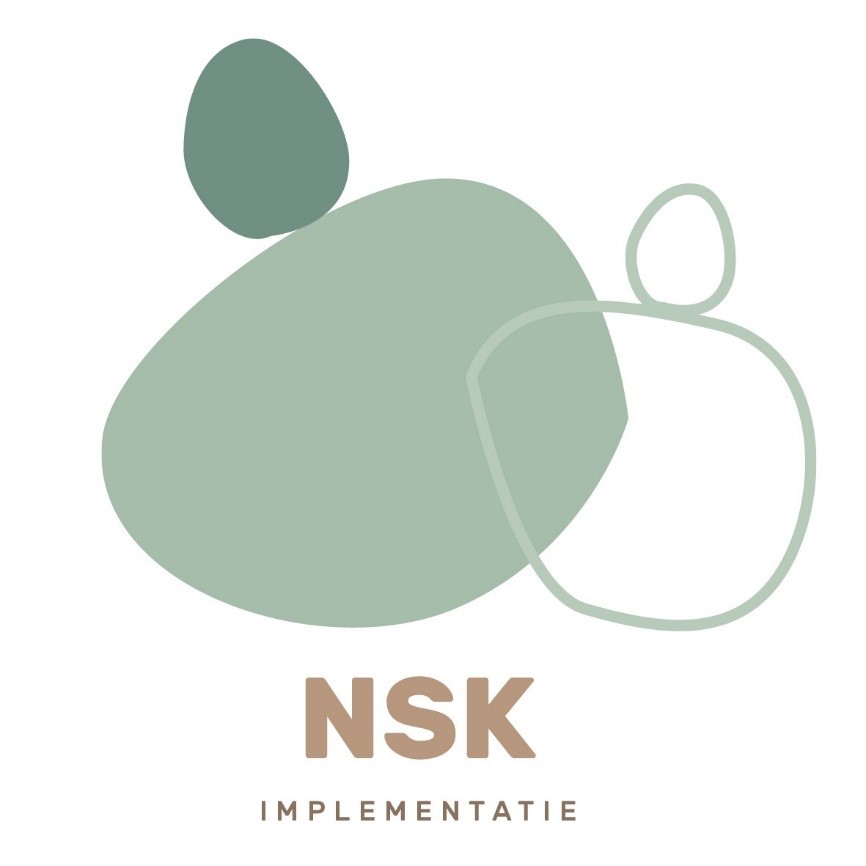

Version: 4.1
Date: June 2023
Author: SCAN research team

Table of content

### Introduction

This implementation strategy provides information regarding the deployment of the Screening Instrument for Child Abuse and Neglect (SCAN). The SCAN screening, along with the SCAN-follow up (SCAN-FU) has been integrated into the electronic health record (EHR) and is available for systems including EPIC, HIX, and Nexus. During the research phase, the SCAN must be mandatorily implemented in the emergency department (ED), but it can also be accessed in both inpatient and outpatient clinical records.

Following several rounds of implementation, this strategy for further implementation has been developed. This document aims to support all clinical ambassadors in ensuring the smooth implementation of the SCAN (SCAN screening + SCAN follow-up).

It is important to note that the implementation of the SCAN within participating hospitals is the responsibility of the clinical ambassadors of that hospital. Any issues arising during or around the implementation process must be reported promptly to the research team to facilitate resolution and prevent recurrence in other centres.

#### Objectives of SCAN

The SCAN screening is a concise and validated instrument for detecting all forms of child abuse. It will be mandatory in the emergency department. Additionally, data will be automatically collected after the implementation process through the SCAN-FU. This anonymised data will contribute to a national healthcare evaluation regarding the detection process. Participating centres will receive an annual national report, which may help to refine the effectiveness of the SCAN at each step of the medical reporting code and provide feedback on how the medical reporting code (KNMG) is used.

###### Core changes

- The SCAN screening is validated and replaces SPUTOVAMO (and its variants).
- Both the SCAN screening and the SCAN-FU are integrated into the EHR.
- In cases of a positive SCAN screening, the SCAN-FU guides the clinician through the steps of reporting code.
- The KNMG reporting code is mandatory for all hospital staff (including nurses and paramedical personnel).
- All information regarding suspected child abuse or unsafe environments is centrally stored within the patient record (protected from patient portal access).
- SCAN screen-positives are automatically compiled into a task list, enabling:
  - Monitoring progress through the reporting code.
  - Efficient preparation of multidisciplinary child abuse team meetings. The task list can serve as an agenda, allowing all aberrant signals to be reviewed (anonymously) with the “Veilig Thuis” organisation.
- The SCAN policy manual serves as a reference document providing additional information about the reporting field, communication, record-keeping, injury documentation, and legal frameworks.

###### Principles and other Key Information

- The SCAN screening must be completed by emergency department nurses for all presenting children, while the SCAN-FU is the physician’s responsibility.
- The SCAN policy manual, which can be tailored regionally, is available in the hospital’s digital protocol database.
- The “index patient” refers to the child, with documentation carried out in the child’s EHR (< 18 years old).
- Although the SCAN and associated procedures are nationally standardised, they can be adapted to the hospital’s specific agreements. Deviations from the standard procedure must be reported to the research team, detailing the points of divergence and their rationale.
- The SCAN and SCAN policy manual are endorsed by the KNMG. Adherence to SCAN procedures ensures compliance with the legal frameworks of the reporting code.
- All necessary information is contained in the SCAN policy manual; clinical ambassadors are advised to familiarise themselves thoroughly with its contents.

### Implementation Timeline

Each participating hospital will establish its own timeline, coordinated by the local team and research team. Broadly, the implementation comprises three phases: pre-implementation, implementation, and post-implementation. The subsequent sections provide further details about each phase.

#### Pre-Implementation

Before SCAN deployment, several steps must be completed:

- Obtain approval from relevant departments (at a minimum, paediatrics and emergency department) and the Board of Directors.
- Appoint clinical ambassador and sign the collaboration agreement.
- Conduct a pre-implementation assessment (baseline measurement) to map the current screening process in the hospital. The research team will provide a format/questionnaire for this purpose.
- Collaborate with the IT department to prioritise the project and establish a timeline.
- Launch the SCAN e-learning module from Augeo within the hospital’s digital learning environment:
  - Identify which professionals must complete the e-learning and in what capacity (mandatory/optional).
  - Free access for hospitals with an Augeo subscription.
  - For hospitals without a subscription, a one-time fee of €400 will apply for the e-learning. This module, designed to familiarise users with the SCAN’s EHR integration, will be available to an unlimited number of users for one year.
- Organise training sessions for and by local SCAN representatives within relevant departments:
  - Determine who will deliver the training. Implementation is typically more successful when driven locally. The research team can assist, subject to agreement with the local team.
- Initiate an internal hospital campaign:
  - Raise awareness through intranet updates, department-specific SCAN information, newsletters, etc.
- Determine a start date:
  - Coordinate with IT and other involved departments.
- Develop an awareness plan to sustain focus on implementation:
  - Consider thematic months, newsletter updates, morning briefings, etc.

To facilitate these steps, consider the following questions:

- Who are the SCAN users?
- How can they be reached for e-learning?
- Do these users belong to a single department/division, or must multiple departments/divisions be involved? Should this be addressed through the hospital’s top-down structure or another approach?

The SCAN research team will provide additional documents to support this process.

The following additional documents are made available:

| **Document** | **Attachment** |
| --- | --- |
| Pocket Card – SCAN | Attachment 1 – Pocket Card |
| Digital Factsheet – SCAN | Attachment 2 – Digital Factsheet SCAN |
| Poster | Attachment 3 – Poster SCAN |
| Training Session Presentation – 45 mins | Attachment 4 – Detailed Presentation |
| Handover Presentation – 10 mins | Attachment 5 – Handover Presentation |
| Case Presentation HIX/EPIC/NEXUS – 10 mins | Attachment 6 – Case Presentation Hix |

The training session presentation provides more information on the purpose of the SCAN, its interface, and usage. This presentation is intended for paediatricians, emergency department physicians, and key users. Additionally, a concise PowerPoint will be provided to guide users through the SCAN using a case example; this can be utilised for practice and to create awareness of the SCAN.

Other informational materials are also available, including standard intranet texts, references to relevant literature, and evidence supporting the SCAN.

#### Implementation

At the start of the implementation phase, thorough preparation is crucial. For unforeseen issues, the research team can always be contacted.

#### Post-Implementation

After three months, an evaluation of the SCAN will take place. Feedback from this evaluation will be shared with the research team to facilitate iterative improvements. During the evaluation period, the SCAN will continue to be used as usual.

Annually, the clinical ambassador is invited to participate in the SCAN group meeting. During these meetings, trends and the annual report are discussed. This provides an opportunity to propose desired modifications to ensure that the SCAN and the reporting field for aberrant signals remain as user-friendly as possible.

We kindly ask you to extract and uploaded every 3–6 months, to gain better insights into the functioning and use of the SCAN. Support for this process will be provided by our data specialists. The anonymisation and upload process has been designed to be as user-friendly as possible, requiring no more than 30 minutes per upload (depending on the number of cases).

The data will also be returned to the hospital in an annual report issued by the LECK. This allows hospitals to see how their performance compares with other hospitals, helping child abuse teams identify areas for process improvement.

The SCAN will remain in use at the centre. Annual SCAN group meetings will be convened to assess the SCAN’s content and processes on a national level and make necessary adjustments. Active representation from project hospitals will be required.

### Checklist

| **Phase** | **Description** | **Responsible Party** | **Deadline** |
| --- | --- | --- | --- |
| **Pre-Implementation** |  |  |  |
|  | Approval from relevant departments and Board of Directors | Clinical ambassador | As soon as possible |
|  | Return signed collaboration agreement | Clinical ambassador | As soon as possible |
|  | Data collection for baseline measurement and submission to research team | Clinical ambassador | As soon as possible |
|  | Preparation for implementation; dissemination of e-learning in Augeo, training sessions, internal hospital campaign, awareness plan, and setting start date | Research team and clinical ambassador | As soon as possible |
|  | Addressing unforeseen problems | Research team and clinical ambassador | As soon as possible |
| **Implementation** |  |  |  |
|  | Start implementation | Clinical ambassador | Date X |
|  | Addressing unforeseen problems | Research team and clinical ambassador | As soon as possible |
| **Post-Implementation** |  |  |  |
|  | Focus group evaluation of initial implementation | Research team and clinical ambassador | X + 12 weeks |
|  | Transition of data to database | Research team and clinical ambassador, ICT of partipating hospital | X + 12 weeks |
|  | Annual project group meetings | Research team and clinical ambassadors | Annually after X |
|  | Addressing unforeseen problems | Research team and clinical ambassador | As soon as possible |

### Contact Information

Paediatrician – Implementation Project Leader
Division of Children, Paediatrics
