## Supplement B for "Towards uniform recognition of child abuse in the Netherlands: implementing the Screening instrument for Child Abuse and Neglect (SCAN)"

|  | Academic | | | | | | Teaching | | | | | General | | | | |  |
| --- | --- | --- | --- | --- | --- | --- | --- | --- | --- | --- | --- | --- | --- | --- | --- | --- | --- |
| MIDI item | **N** | **Mean (SD)** | **disagree / totally disagree** | **neutral** | | **agree / totally agree** | **N** | **Mean (SD)** | **disagree / totally disagree** | **neutral** | **agree / totally agree** | **N** | **Mean (SD)** | **disagree / totally disagree** | **neutral** | **agree / totally agree** | **ANOVA** |
| 1.Procedural clarity | 60 | 3.63 (0.66) | 6.7 | | 26.7 | 66.6 | 98 | 3.77 (0.76) | 6.2 | 15.3 | 78.5 | 34 | 3.82 (0.57) | 6.3 | 17.7 | 76 | 0.37 |
| 2.Completeness | 58 | 3.53 (0.71) | 8.6 | | 32.8 | 58.6 | 95 | 3.72 (0.61) | 4.2 | 24.2 | 71.6 | 32 | 3.65 (0.65) | 6.3 | 25 | 68.7 | 0.25 |
| 3.Complexity* | 58 | 3.52 (0.92) | 55.2 | | 31 | 13.8 | 94 | 3.85 (0.95) | 69.2 | 21.3 | 9.6 | 32 | 3.72 (0.81) | 62.5 | 31.3 | 6.3 | 0.09 |
| 4.Compatibility | 58 | 3.43 (0.82) | 10.3 | | 36.2 | 53.4 | 93 | 3.67 (0.70) | 6.5 | 23.7 | 69.9 | 32 | 3.56 (0.76) | 15.6 | 18.8 | 65.6 | 0.15 |
| 5.Relevance to patient | 57 | 3.65 (0.61) | 3.5 | | 31.6 | 64.9 | 92 | 3.87 (0.65) | 4.4 | 12 | 83.7 | 31 | 3.65 (0.66) | 9.7 | 16.1 | 74.2 | 0.07 |
| 6a.Personal advantages/disadvantages - actions | 55 | 3.27 (0.89) | 20 | | 36.4 | 43.7 | 83 | 3.58 (0.77) | 9.6 | 30.1 | 60.2 | 30 | 3.03 (0.81) | 16.7 | 56.7 | 26.7 | <0.01 |
| 6b. Personal advantages/disadvantages - documentation | 55 | 3.47 (0.63) | 7.3 | | 38.2 | 54.5 | 83 | 3.40 (0.73) | 8.4 | 45.8 | 45.8 | 29 | 3.17 (0.71) | 17.2 | 48.3 | 34.5 | 0.17 |
| 7a.Outcome expectations - recognition | 55 | 3.85 (0.71) | 3.6 | | 21.8 | 74.5 | 81 | 3.77 (0.68) | 4.9 | 22.2 | 72.8 | 29 | 3.79 (0.72) | 6.8 | 24.1 | 69.1 | 0.68 |
| 7b.Outcome expectations – expectance | 54 | 3.22 (0.66) | 9.3 | | 63 | 27.8 | 80 | 3.36 (0.68) | 6.3 | 56.3 | 37.5 | 29 | 3.17 (0.76) | 20.7 | 41.4 | 37.9 | 0.33 |
| 8.Perception of responsibility | 54 | 4.20 (0.71) | 1.9 | | 11.1 | 87.1 | 80 | 4.31 (0.67) | 1.3 | 7.5 | 91.3 | 29 | 4.10 (0.94) | 6.8 | 6.9 | 86.3 | 0.39 |
| 9.Social support | 54 | 4.19 (0.65) | 13 | | 55.6 | 31.5 | 80 | 4.00 (0.69) | 18.8 | 60 | 21.3 | 29 | 3.93 (0.65) | 17.2 | 69 | 13.8 | 0.17 |
| 10a.Self-efficacy – SCAN screening | 53 | 4.02 (0.60) | 0 | | 17 | 83.1 | 80 | 4.16 (0.67) | 1.3 | 7.5 | 91.2 | 29 | 4.14 (0.44) | 0 | 3.4 | 96.5 | 0.40 |
| 10b.Self-efficacy – SCAN follow-up | 53 | 3.79 (0.72) | 3.8 | | 26.4 | 69.8 | 80 | 3.66 (0.67) | 5.1 | 26.3 | 68.8 | 29 | 3.52 (0.87) | 6.8 | 41.4 | 51.7 | 0.25 |
| 11.Knowledge | 53 | 3.87 (0.65) | 1.9 | | 22.6 | 75.5 | 79 | 3.90 (0.69) | 5.1 | 10.1 | 84.8 | 29 | 3.76 (0.64) | 3.4 | 24.1 | 72.4 | 0.63 |
| 12.Formal ratification by management^#^ | 52 | 1.85 (0.36) | 15.4 | |  | 84.6 | 78 | 1.85 (0.36) | 15.4 |  | 84.6 | 29 | 1.93 (0.26) | 6.9 |  | 93.1 | 0.49 |
| 13.Staff capacity | 52 | 3.52 (0.78) | 9.6 | | 36.5 | 53.9 | 76 | 3.67 (0.68) | 6.6 | 21.1 | 72.3 | 29 | 3.66 (0.48) | 0 | 34.5 | 65.5 | 0.45 |
| 14.Availablility of materials and resources | 52 | 3.60 (0.57) | 1.9 | | 38.5 | 59.6 | 79 | 3.74 (0.70) | 5.2 | 21.1 | 73.7 | 29 | 3.83 (0.38) | 0 | 17.2 | 82.8 | 0.22 |
| 15.Unsettled organisation^#^* | 53 | 1.43 (0.50) | 56.6 | |  | 43.3 | 76 | 1.59 (0.50) | 40.8 |  | 59.2 | 28 | 1.64 (0.49) | 35.7 |  | 64.3 | 0.11 |
| 16.Acces to information on SCAN tools use | 53 | 3.85 (0.66) | 1.9 | | 24.5 | 73.6 | 76 | 3.84 (0.61) | 2.6 | 15.8 | 81.6 | 28 | 3.61 (0.57) | 3.6 | 32.1 | 64.3 | 0.19 |
| 17.Legislations and regulations | 51 | 3.76 (0.71) | 3.9 | | 27.5 | 68.7 | 76 | 4.01 (0.58) | 0 | 15.8 | 84.2 | 28 | 3.71 (0.66) | 7.1 | 17.9 | 75 | 0.04 |

**Supplement table 1: Subgroup analysis hospital type**

|  | 0-5 years | | | | | 6-15 years | | | | | >15 years | | | | |  |
| --- | --- | --- | --- | --- | --- | --- | --- | --- | --- | --- | --- | --- | --- | --- | --- | --- |
| MIDI item | **N** | **Mean (SD)** | **disagree / totally disagree** | **neutral** | **agree / totally agree** | **N** | **Mean (SD)** | **disagree / totally disagree** | **neutral** | **agree / totally agree** | **N** | **Mean (SD)** | **disagree / totally disagree** | **neutral** | **agree / totally agree** | **ANOVA** |
| 1.Procedural clarity | 59 | 3.76 (0.77) | 6.8 | 9.1 | 84.1 | 87 | 3.82 (0.60) | 4.1 | 13.7 | 82.2 | 67 | 3.70 (0.71) | 5.4 | 17.9 | 76.8 | 0.59 |
| 2.Completeness | 53 | 3.66 (0.78) | 11.4 | 15.9 | 72.7 | 87 | 3.69 (0.60) | 4.1 | 23.3 | 72.6 | 65 | 3.60 (0.66) | 7.1 | 25 | 67.9 | 0.71 |
| 3.Complexity* | 52 | 3.85 (0.85) | 68.2 | 25 | 6.8 | 87 | 3.71 (0.94) | 67.1 | 19.2 | 13.7 | 65 | 3.74 (0.92) | 67.8 | 21.4 | 10.7 | 0.70 |
| 4.Compatibility | 52 | 3.62 (0.72) | 6.8 | 20.5 | 72.8 | 86 | 3.57 (0.73) | 9.3 | 24.7 | 65.9 | 65 | 3.54 (0.79) | 10.7 | 28.6 | 60.7 | 0.86 |
| 5.Relevance to patient | 50 | 3.76 (0.74) | 6.8 | 13.6 | 79.5 | 86 | 3.81 (0.58) | 2.7 | 21.9 | 75.3 | 64 | 3.75 (0.64) | 5.4 | 16.1 | 78.6 | 0.80 |
| 6a.Personal advantages/disadvantages - actions | 45 | 3.58 (0.89) | 13.6 | 22.7 | 63.6 | 82 | 3.35 (0.81) | 16.4 | 34.2 | 49.3 | 61 | 3.33 (0.81) | 10.7 | 46.4 | 42.9 | 0.25 |
| 6b. Personal advantages/disadvantages - documentation | 45 | 3.33 (0.83) | 15.9 | 36.4 | 47.8 | 81 | 3.43 (0.71) | 9.6 | 41.1 | 49.3 | 60 | 3.42 (0.70) | 7.1 | 42.9 | 50 | 0.76 |
| 7a.Outcome expectations - recognition | 45 | 4.00 (0.67) | 2.3 | 15.9 | 81.9 | 80 | 3.74 (0.65) | 5.5 | 21.9 | 72.6 | 59 | 3.76 (0.80) | 5.4 | 19.6 | 75 | 0.12 |
| 7b.Outcome expectations – expectance | 45 | 3.29 (0.70) | 9.1 | 59.1 | 31.8 | 78 | 3.28 (0.66) | 9.6 | 56.2 | 34.3 | 58 | 3.33 (0.74) | 10.7 | 51.8 | 37.5 | 0.93 |
| 8.Perception of responsibility | 45 | 4.22 (0.70) | 2.3 | 9.1 | 88.7 | 78 | 4.32 (0.67) | 1.4 | 6.8 | 91.7 | 58 | 4.21 (0.79) | 3.6 | 5.4 | 91.1 | 0.61 |
| 9.Social support | 45 | 4.27 (0.75) | 2.3 | 11.4 | 86.4 | 78 | 3.92 (0.68) | 4.1 | 15.1 | 80.9 | 58 | 4.00 (0.56) | 0 | 14.3 | 85.7 | 0.02 |
| 10a.Self-efficacy – SCAN screening | 44 | 4.16 (0.65) | 0 | 13.6 | 86.3 | 78 | 4.12 (0.62) | 1.4 | 5.5 | 93.1 | 58 | 4.09 (0.51) | 0 | 7.1 | 92.9 | 0.83 |
| 10b.Self-efficacy – SCAN follow-up | 44 | 3.77 (0.68) | 4.5 | 22.7 | 72.7 | 78 | 3.73 (0.72) | 4.1 | 24.7 | 71.2 | 58 | 3.57 (0.75) | 5.4 | 37.5 | 57.1 | 0.29 |
| 11.Knowledge | 44 | 3.86 (0.59) | 4.5 | 11.4 | 84.1 | 78 | 3.85 (0.72) | 5.5 | 13.7 | 80.9 | 57 | 3.91 (0.58) | 0 | 19.6 | 80.4 | 0.84 |
| 12.Formal ratification by management^#^ | 44 | 1.89 (0.32) | 11.4 |  | 88.6 | 76 | 1.87 (0.34) | 12.3 |  | 87.7 | 57 | 1.86 (0.35) | 14.3 |  | 85.7 | 0.92 |
| 13.Staff capacity | 44 | 3.66 (0.65) | 4.5 | 29.5 | 65.9 | 75 | 3.55 (0.72) | 8.2 | 28.8 | 63 | 56 | 3.70 (0.66) | 5.4 | 25 | 69.7 | 0.43 |
| 14.Availablility of materials and resources | 44 | 3.73 (0.59) | 2.3 | 27.3 | 70.4 | 75 | 3.60 (0.68) | 5.5 | 30.1 | 64.3 | 56 | 3.84 (0.53) | 1.8 | 17.9 | 80.4 | 0.09 |
| 15.Unsettled organisation^#^* | 44 | 1.66 (0.48) | 65.9 |  | 34.1 | 75 | 1.49 (0.50) | 50.7 |  | 49.3 | 56 | 1.55 (0.50) | 55.4 |  | 44.6 | 0.22 |
| 16.Acces to information on SCAN tools use | 44 | 3.98 (0.51) | 0 | 13.6 | 86.4 | 75 | 3.76 (0.59) | 1.4 | 24.7 | 73.9 | 56 | 3.77 (0.69) | 5.4 | 21.4 | 73.2 | 0.13 |
| 17.Legislations and regulations | 44 | 3.84 (0.57) | 2.3 | 18.2 | 79.5 | 73 | 4.01 (0.61) | 0 | 17.8 | 82.2 | 56 | 3.79 (0.71) | 5.4 | 21.4 | 73.2 | 0.11 |

**Supplement table 2: Subgroup analysis work experience**
